# “Prevalence and Determinants of Betel Quid Dependence in Rural Bangladesh: A Cross-Sectional Analysis of Sociodemographic and Behavioral Factors“

**DOI:** 10.1101/2025.06.24.25330179

**Authors:** Md. Rizwanul Karim, Shahnaz Akhter, Taibah Tanzil, Tahmid Sajid, Shahida Afnan, Taslima Zannat

## Abstract

**Background:** Areca nut, the main component of betel quid (BQ) and the fourth most-used psychoactive substance worldwide, is consumed by over 600 million people, primarily in South Asia, and is a recognized carcinogen linked to serious health risks, especially oral conditions such as precancerous lesions and oral cancer. This study aimed to assess the dependency among BQ users and identify the key sociodemographic and behavioral predictors that significantly influence this dependency.

**Method:** This cross-sectional study was carried out between January and June 2023 on a randomly selected sample of 400 individuals from Bhaluka Upazila in the Mymensingh district of Bangladesh. A structured questionnaire collected data on sociodemographics, BQ consumption, chewing motivations, quitting intentions, and dependency levels. Data analysis was conducted using SPSS and Jamovi, employing confirmatory factor analysis, structural equation modeling, and binary logistic regression.

**Results:** The study used a seven-item, three-factor adapted Betel Quid Dependency Scale with good reliability (Cronbach’s α = 0.83). The study revealed that approximately half of the participants (48.5%) were BQ-dependent, with a higher prevalence observed among females. Key predictors of dependence included education levels, expenditures on BQ, tobacco with BQ, and unsuccessful attempts to quit. Among BQ-tobacco users, more than half of the females primarily consumed smokeless tobacco, and the rate of failed quit attempts among females was more than twice that of males. The reasons for chewing BQ were found to predict dependency, while the intention to quit BQ had an opposing effect.

**Conclusion:** The study validated a shortened version of the Betel Quid Dependence Scale and identified key sociodemographic and behavioral determinants of dependence, including education level, intention to quit, previous quit attempts, and expenditure on betel quid. It recommends implementing health education campaigns, providing behavioral support, strengthening anti-tobacco policies, and designing targeted interventions for high-risk populations to reduce BQ dependency.

## Background

The term “Betel quid” refers to a diverse mixture of ingredients and cultural practices, commonly known as *“pan*” or “*paan”*. It typically consists of betel leaf, areca nut, and slaked lime, often combined with tobacco and regional spices or sweeteners to enhance the flavor. Globally, an estimated 600 million people chew betel quid (BQ), with dependency rates ranging from 20 to 90% [1]. Areca nut, the fourth most consumed psychoactive substance after tobacco, alcohol, and caffeine, is used by 10% to 20% of the global population as part of BQ. This practice is widespread in South and Southeast Asia, the Asia–Pacific region, and among migrant communities in Africa, North America, and Europe [2].

In Bangladesh, BQ, locally known as “*pan,”* holds significant cultural importance, particularly in rural areas. Approximately 75% of users combine it with tobacco products like “*Zarda*”, “*Sadapata*/*Pata*”, or “*Gul*.” Among Bangladeshi adults, 18.7% use BQ with tobacco, with a higher prevalence among women (23.0%) than men (14.3%) [3, 4, 5]. Firmly rooted in social traditions, betel quid (BQ) is commonly shared at gatherings and is thought to support digestion. Key factors affecting BQ usage include gender, age, education, occupation, and economic status [6]. Betel quid chewing is strongly associated with severe oral health issues, including oral cancer, which is the second leading cause of cancer mortality among men in Southeast Asia and accounts for one-third of global cancer cases [7]. A meta-analysis found an adjusted relative risk of 7.9 for oral cancer among BQ users, with nearly half of all cases in the Indian subcontinent and Taiwan attributed to its use [ 8]. The International Agency for Research on Cancer (IARC) classifies areca nut and BQ (even without tobacco) as carcinogenic [9,10]. Regular consumption damages oral tissues, causing tooth attrition, periodontitis, and lesions such as betel mucosa, increasing cancer risk [11, 12]. Betel quid consumption triggers oral and pharyngeal cancer-related pathways and causes significant genetic damage, with the highest risk observed in those who also smoke [13, 14]. It also disrupts the oral microbiome, contributing to oral diseases and cancer [15,16]. It is linked to conditions such as oral submucous fibrosis, leukoplakia, and systemic diseases like obesity, cardiovascular disease, and diabetes [17, 18, 19, 20]. Long-term betel quid use was found to increase the risk of significant liver fibrosis in people with metabolic syndrome (MetS), while no similar risk was seen in those without MetS [21]. Additionally, habitual BQ use and spitting have been linked to tuberculosis transmission, underscoring the need for further research [22].

Betel quid’s addictive and harmful effects stem from two distinct mechanisms: toxic high-weight compounds damage oral tissues, while low-weight components interact with multiple brain chemicals (GABA, glutamate, dopamine, serotonin) to create addiction. This dual action simultaneously causes physical harm and psychological dependence [23]. Neuroimaging studies reveal that BQ chewing activates brain regions associated with reward, movement, and vision, suggesting that dependency alters brain connectivity and reinforces addictive behaviors [24]. Areca nut, particularly when combined with tobacco or smoking, significantly increases the risk of dependence syndrome [25]. Areca nuts contain alkaloids, primarily arecoline, which stimulate the central and autonomic nervous systems, elevating noradrenaline and acetylcholine levels. Dependence manifests through cognitive, behavioral, and physiological symptoms, including loss of control and continued use despite adverse effects. Short-term effects of arecoline include increased facial skin temperature, palpitations, and sweating, while long-term use is associated with heightened alertness, attention, and appetite suppression [26, 27]. Betel quid intoxication increases heart rate and facial temperature, and affects psychological perception, including time perception, arousal, and cognitive function; however, it does not significantly impact working memory or reaction time [28]. Despite these findings, research on areca nut dependence remains limited, focusing mainly on withdrawal symptoms during abstinence. Dependency diagnosis requires symptoms such as tolerance, withdrawal, loss of control, and cravings [29, 30]. Regular BQ users often exhibit tolerance and withdrawal symptoms like nicotine addiction [30]. A recent systematic review revealed that dependent BQ users struggled to quit BQ due to habituation, addiction, and withdrawal symptoms. Additionally, some social BQ chewers in Southeast Asia were reluctant to quit BQ, viewing areca nut (AN) as a cultural and social identifier, with peer influence reinforcing their habit [31]. Individuals meeting DSM-5 criteria for BQ use disorder, such as failed quit attempts, neglect of responsibilities, social conflicts, and continued use despite health risks, face a higher risk of developing oral squamous cell carcinoma (OSCC) [32].

Social learning theories suggest that substance use behaviors are influenced by social, cultural, behavioral, and physiological factors. A cross-sectional study in Guam examined the Reasons for Betel-quid Chewing Scale (RBCS), a 10-item measure adapted from smoking motivation scales. The study identified stimulation as the most common reason for use, followed by reinforcement and social/cultural factors [33, 34]. Despite its health risks, limited research has explored willingness to quit, with betel quid dependence patterns resembling those of tobacco use, indicating a need for similar cessation strategies [30, 35]. However, A recent study introduced “betel years” as a new measure to quantify lifetime betel quid exposure, accounting for duration and additives like slaked lime and tobacco. Analysis linked this measure to cellular changes and oral precancerous signs, offering a standardized tool for risk assessment similar to “pack years“ in smoking [36]. The Betel Quid Dependence Scale (BQDS), validated in Taiwan, is a self-assessment tool with high internal consistency across three dimensions: “physical and psychological urgent need,” “increasing dose, “and” maladaptive use. “A study found that 42.2% of male prisoners met DSM-IV criteria for dependent use, with the BQDS achieving a predictive accuracy of 99.3% at a cut-off score of 4 [37]. The scale was also validated in Guam through community outreach [38].

Areca nut use is more prevalent among men, who are more likely to recognize its negative health and economic impacts than women [39, 40]. In Taiwan, men exhibit higher rates of betel quid use, while women show lower cessation rates, influenced by education and ethnicity. Both initiation and cessation are significantly affected by various factors, while smoking, occupation, and alcohol use primarily impact initiation [41]. The Global Adult Tobacco Survey (GATS) in India found a 40% decline in BQ use among young males who also used tobacco, whereas the usage among females remained unchanged. Tobacco-laced betel quid is more common in lower-income groups; however, non-tobacco use is prevalent among those with moderate education, highlighting the need for targeted public health interventions [42].

Areca nut addiction, often overlooked compared to tobacco, is driven by social norms, stress, and limited awareness. Despite its addictive potential, few cessation programs exist, and many are adapted from tobacco interventions. Barriers to quitting include sociocultural influences, behavioral factors, and accessibility to betel quid. Current research explores behavioral interventions and pharmacological therapies [43]. Given its severe health risks, betel quid consumption requires dedicated prevention strategies beyond general tobacco control measures [44, 45, 46]. The issue is exacerbated by the emergence of new betel-based products, aggressive promotional strategies, and deeply ingrained cultural practices, particularly in South Asia, where individuals often begin chewing during adolescence. Among migrant populations, the habit persists due to low health literacy and limited access to dental care [46]. The research gap lies in the limited understanding of sociodemographic and behavioral determinants of BQ dependence in rural populations, particularly in low-resource settings, and the lack of evidence-based cessation programs tailored to address its unique cultural and addictive dynamics [47].

Examining betel quid (BQ) dependence in rural communities is crucial, given its higher prevalence, strong cultural ties, and the influence of educational, occupational, and economic disparities. Additionally, limited access to healthcare and low levels of awareness in these settings pose significant challenges to intervention efforts, underscoring the need for tailored and context-specific strategies. This study aims to assess the prevalence of BQ dependence in rural communities and identify sociodemographic and behavioral factors involved, providing insights for targeted and effective public health interventions.

### Conceptual framework

An empirically grounded relationship model of betel quid dependence was conceptualized, integrating insights from multiple social science theories through a comprehensive literature review [Fig.1].

**Fig. 1.**
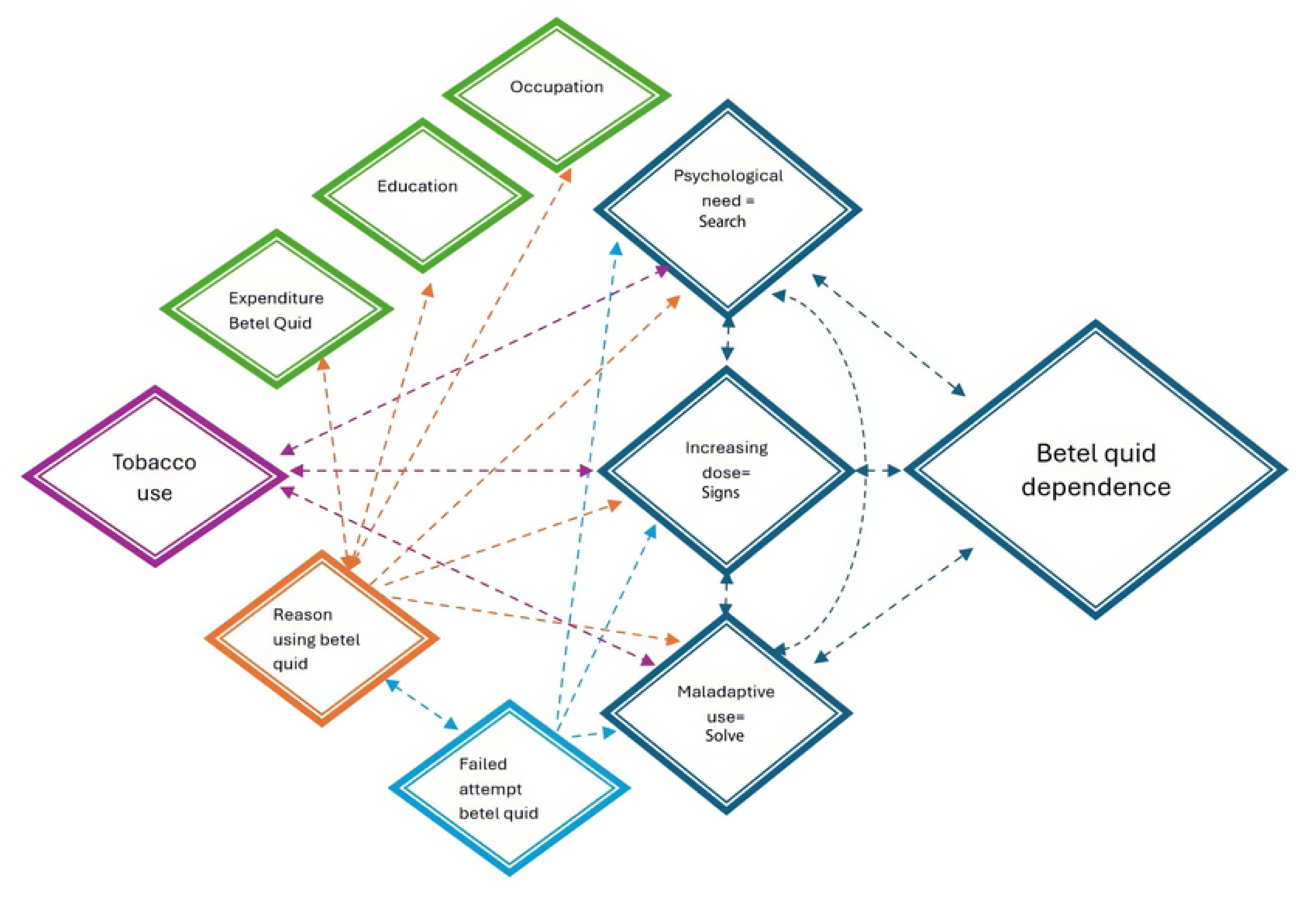
A hypothesized conceptual framework of factors contributing to Betel quid dependence

The conceptual framework presented in the figure provides a comprehensive model for understanding betel quid dependence, integrating socioeconomic, psychological, and social factors. Socioeconomic influences such as occupation, education, and expenditure align with the “Strain Theory,” suggesting that individuals may turn to betel quid as a coping mechanism for economic stress or limited opportunities. Psychological needs and maladaptive use are explained by “Cognitive-Behavioral Theory,” which highlights how individuals may rely on betel quid to manage stress or emotional distress, leading to cognitive distortions and reinforcing dependence. Social influences, including cultural norms and peer behavior, are addressed by “Social Learning Theory,” which posits that individuals learn and adopt betel quid use through observation and social reinforcement. The dose–response relationship and increasing dependence are linked to “Biosocial Theories,” emphasizing the interaction between biological factors (e.g., arecoline in betel quid, tobacco with betel quid) and environmental influences. Finally, “Rational Choice Theories” considers individuals’ decision-making processes. Failed attempts to quit may result from weighing perceived benefits (e.g., stress relief) against costs (e.g., health risks), often influenced by addiction and social pressures. Together, these theories offer a comprehensive framework for understanding betel quid dependence and its multifaceted drivers.

### Ethics approval and consent to participate

The Patuakhali Medical College Research Ethics Committee provided ethical approval for this survey (Reference no: PkMC–REC–23–01–S12). All ethical issues involved in this study, including privacy, confidentiality, and anonymity, were carefully maintained following the Helsinki Declaration. Before beginning the data collection procedure, respondents were given a brief overview of the study’s goals and objectives. They were fully informed of their right to engage or reject participation in the study. Their signed consent was then acquired using a separate consent form affixed with the semi-structured interviewer-administered survey questionnaire. The participants in the study were assured that any personally identifiable information they provided would be kept private and anonymous and that the study’s findings would only be shared and published for the public interest.

## Method

### Study design and sample size determination

This cross-sectional study was conducted in the rural communities of four of the eleven Unions in Bhaluka Upazila, Mymensingh district, Bangladesh, between January and June 2023. The research employed a simple random sampling method to ensure the collection of representative data. The calculated sample size was 370, determined using the “Epitools” online sample size calculator (https://epitools.ausvet.com.au/samplesize), based on a 95% confidence level, 5% margin of error, and an assumed prevalence of betel quid dependency at 40.5% [16]. The intended sample size was 400 respondents, considering possible non-response and missing data.

### Sampling

In this study, samples were collected randomly from rural communities across four of the eleven Unions in the Bhaluka Upazila, located in the Mymensingh district. A total of (50*2 Wards = 100*4Unions = 400) four hundred samples were collected from the four randomly chosen Unions in Bhaluka Upazila [Fifty samples were taken from each of two randomly selected Wards out of the nine Wards* in each Union (*Lowest local-government administrative tiers consist of approximately 1200 families comprising 6000 population)]. The sampling frame for this study was prepared from the Bangladesh Bureau of Statistics (BBS) register used in the Global Adult Tobacco Survey (GATS) 2017, along with the NID register from the Bhaluka Upazila Administration. Before data collection began, the Bhaluka Upazilla Administration received a letter describing the study’s goals and objectives. The research team then scheduled a study protocol discussion session for the selected Unions (Birunia, Uthura, Mallikbari, and Dhitpur) and Ward authorities, coordinating with the Bhaluka-Upazila Administrator’s office. The study included households with male and/or female participants aged 18 to 60 who consumed at least one betel quid daily over the past year. If no adult betel quid user was found in a randomly selected household at the Ward level, the sample was taken from a nearby adjacent household or the one next to it. Individuals aged 18 to 60, without severe illness or psychiatric disorders, who consented to participate, were interviewed. All eligible and available individuals were approached, irrespective of gender or social status.

### Data collection method

The questionnaire was initially created in English, then translated into Bengali, and later back-translated by two independent multilingual translators to ensure reliability and reduce response bias. The interviewer-administered structured questionnaire was pretested among 30 respondents of both sexes in an adjacent upazila (Gofargaon Upazila) to ensure its relevance, clarity, and appropriateness. After the pre-testing, any irrelevant or problematic components were identified and corrected. In-person interviews were carried out following the acquisition of formal consent from the research participants. Participants took about 30 minutes to complete the interview, ensuring complete visual and verbal privacy. A comprehensive review of inconsistencies and missing data confirmed that all 400 respondents had answered every question on the survey questionnaire.

### Data collection instrument

The data collection instrument was divided into six sections: sociodemographic information, personal betel quid (BQ) and or tobacco habits, patterns of BQ use, reasons for chewing BQ, attempts to quit BQ consumption, and dependence on BQ (which includes physical and psychological needs, maladaptive use, and increased dosage).

Data were collected for each block of the conceptual framework. Section 1 of the questionnaire focused on socioeconomic parameters such as age, gender, education, employment, marital status, and income. Section 2 contained questions related to BQ consumption, including the duration of use, age at which use began, substances consumed with BQ, and the quantity and frequency of BQ consumed. In addition to BQ use characteristics, Sect. 3 addressed issues related to tobacco use, such as smoking status, use of smokeless tobacco (SLT), types of tobacco products used, and frequency of use, following the questions from the GATS-Bangladesh and WHO-STEPS Noncommunicable Disease Risk Factor Survey. Section 4 collected data on individual motivations for chewing BQ, while Section. 5 included questions about quitting behavior. Finally, Sect. 6 assessed dependence on betel quid.

### The Reason for Betel Quid Chewing Scale (RBCS)

The Reason for Betel Quid Chewing Scale RBCS is a 10-item, five-point Likert scale (0 = “not important” to 4 = “extremely important”) designed to assess the reasons for chewing betel quid. The three main reasons for chewing betel quid are reinforcement, social and cultural factors, and stimulation. The whole RBCS score and the three subscales demonstrated strong internal consistency, with the overall 10-item RBCS having a Cronbach’s alpha of 0.88 [34].

### Intention to quit betel quid

A five-item, five-point Likert scale was used to collect information on the intention to quit chewing betel quid. Each item inquiry featured five response possibilities (“No, certainly not” = 1, “No, probably not” = 2, “Don’t know/undecided” = 3, “Yes, possibly” = 4, “Yes, definitely” = 5) [35]. According to this study, this single component measure demonstrated outstanding reliability with Cronbach’s alpha of 0.92, as well as adequate convergent and discriminant validity with Average Variance Extracted (AVE) = 0.76 > 0.50, Composite Reliability (CR) = 0.93 > 0.70.

### The Betel Quid Dependence Scale BQDS

In this study, Betel quid dependency was measured, reported, and interpreted using an adapted and validated version of BQDS. The original Betel Quid Dependence Scale BQDS, which has 16 items, was used to measure Betel Quid dependence. Each item had a binary response: “yes” or “no”. The score for “yes” was one, while the score for “no” was zero. The BQDS had a total score that could vary from 0 to 16, and users of betel quid who met three or more of the specific criteria for dependency were considered dependent users. The original BQDS features a three-factor structure with” physical and psychological urgent need,” “increasing dose,” and “maladaptive use, “accounting for 61.2% of the total variance. It also had strong internal consistency (Cronbach’s α = 0.921). The receiver operating characteristic (ROC) curve indicated that the sixteen-item BQDS had an optimal cut-off score of 4, the optimal sensitivity was 0.926, and the optimal specificity was 0.977. The predictive accuracy of the BQDS was up to 99.3% [37, 38].,

### Adaptation and validation of the betel quid dependency scale Exploratory factor analysis

Because the Betel Quid Dependence Scale had not previously been utilized or validated in the Bengali population, we translated and validated the BQDS using an appropriate statistical approach.

Principal factors were extracted with varimax rotation using jamovi-2.6.17.0 on 16 items from the BQDS for a sample of 400 respondents. Principal components extraction was used before principal factors extraction to estimate the number of factors, detect the presence of outliers, assess the absence of multicollinearity, and verify the factorability of the correlation matrices. The Kaiser–Meyer–Olkin (KMO) value of 0.73 indicates an acceptable level of sampling adequacy, as it exceeds the recommended threshold of 0.60 [48]. A higher KMO value indicates that the data are suitable for factor analysis, as it demonstrates sufficient correlations among variables. In addition, Bartlett’s Test of Sphericity achieved statistical significance, indicating that the correlation matrix is not an identity matrix. This supports the assumption that meaningful factors exist within the data, justifying the application of factor analysis [49]. Using Cattell’s scree test, a clear break was observed after the third component, which informed the decision to retain three components for further analysis [50]. This distinction was further supported by parallel analysis. The three-component solution accounted for 71.1% of the variance, with Component 1 contributing 27.27%, Component 2 contributing 24.28%, and Component 3 contributing 19.55%. With a cutoff of 0.40 for the inclusion of a variable in interpreting a factor, 9 of 16 variables did not load on any factor. Failure of numerous variables to load on a factor reflects the heterogeneity of items on the BQDS. Loadings of variables on factors, uniqueness, and percentages of variance are shown in [Table 1].

**Table 1.** Exploratory Factor Analysis: Factor Loadings, explained variance, and reliability. [Scale items with loadings <.40 were not retained].

| Betel quid dependency scale | Factor |  |  | KMO | Variance | Cronbach's |  |
| --- | --- | --- | --- | --- | --- | --- | --- |
| Retained items | 1 | 2 | 3 | Uniqueness MSA | % | $\alpha$ | |
| BQDS3: Whenever you want to chew betel quid but not available, would you spend much time to find it? | 0.93 |  |  | 0.03 | .66 | 27.27 | .96 |
| BQDS4: Whenever you want to chew betel quid but not available, would you take extra steps and travel a great distance trying to buy it? | 0.89 |  |  | 0.11 | .66 |  |  |
| BQDS7: Have you experienced depression or drowsiness after you reduce or completely stop chewing betel quid? |  | 0.75 |  | 0.37 | .83 | 24.28 | .80 |
| BQDS5: Have you felt agitated, irritated, or anxious after you reduce or completely stop chewing betel quid? |  | 0.74 |  | 0.37 | .83 |  |  |
| BQDS6: Have you experienced difficulties concentrating or focusing after you reduce or completely stop chewing betel quid? |  | 0.59 |  | 0.49 | .91 |  |  |
| BQDS11: Have you felt the need to increase the number of betel quid chewing periodically to achieve a pleasant or refreshing effect? |  |  | 0.99 | 0.00 | .64 | 19.55 | .69 |
| BQDS16: Have you reduced or given up any of your social, work, or leisure activities because of betel quid chewing? |  |  | 0.54 | 0.66 | .67 |  |  |
| Extraction Method: Parallel analysis based on factor analysis | Search | Signs | Solve |  | Overall | Total | Overall |
| Rotation Method: Oblimin with Kaiser Normalization. |  |  |  |  | .73 | 71.1 | .83 |
| Bartlett's Test of Sphericity: $p = .000$ | | | | | | | |

Variables are ordered and grouped by size of loading to facilitate interpretation [Zeros replace loadings under 0.40]. The rotated solution revealed the presence of a simple structure, with three components showing no cross-loadings and all variables loading substantially on only one component [51]. The original version of the Betel quid dependency scale featured three elements; the first factor, “physical and psychological need,” had seven items, the second factor, “increasing dose,” had five items, and the third factor,” maladaptive use, “had four variables. On the contrary, the retained seven items, constituting three elements, obtained from the rotational solution based on the present data, were designated “search,” “signs,” and “solve,” respectively. Thus, the three factors on the BQDS for this sample are: search (e.g., “Whenever you want to chew betel quid but not available, would you spend much time to find it?”, “Whenever you want to chew betel quid but are not available, would you take extra steps and travel a great distance trying to buy it?” sign (e.g., “Have you felt agitated, irritated, or anxious after you reduce or completely stop chewing betel quid?”, “Have you experienced difficulty in concentrating or focusing after you reduce or completely stop chewing betel quid?”, “Have you experienced depression or drowsiness after you reduce or completely stop chewing betel quid?” and solve (e.g., “Have you felt the need to increase the amount of betel quid chewing periodically to achieve a pleasant or refreshing effect?”, “Have you reduced or given up any of your social, work, or leisure activities because of betel quid chewing?”) [Table 1].

### Confirmatory factor analysis

Confirmatory factor analysis was employed to specify and test the model fit based on the factorial model generated by exploratory factor analysis [52] [Fig.2]. The model was re-evaluated for theoretical validity since the EFA yielded a three-factor solution. Structural and cultural validity was estimated by evaluating standardized factor loading and fit indices to assess the consistency of the factorial structure. The most often used fitting function in applied CFA and SEM research is maximum likelihood (ML). Certain fundamental assumptions, such as a high sample size, factor indicators measured on continuous scales, and a multivariate normal distribution, may render ML ineffective as an estimator in certain situations. If nonnormality is severe, ML will provide inaccurate parameter estimates; instead, estimators such as WLSMV and ULS should be used [53].

**Fig. 2.**
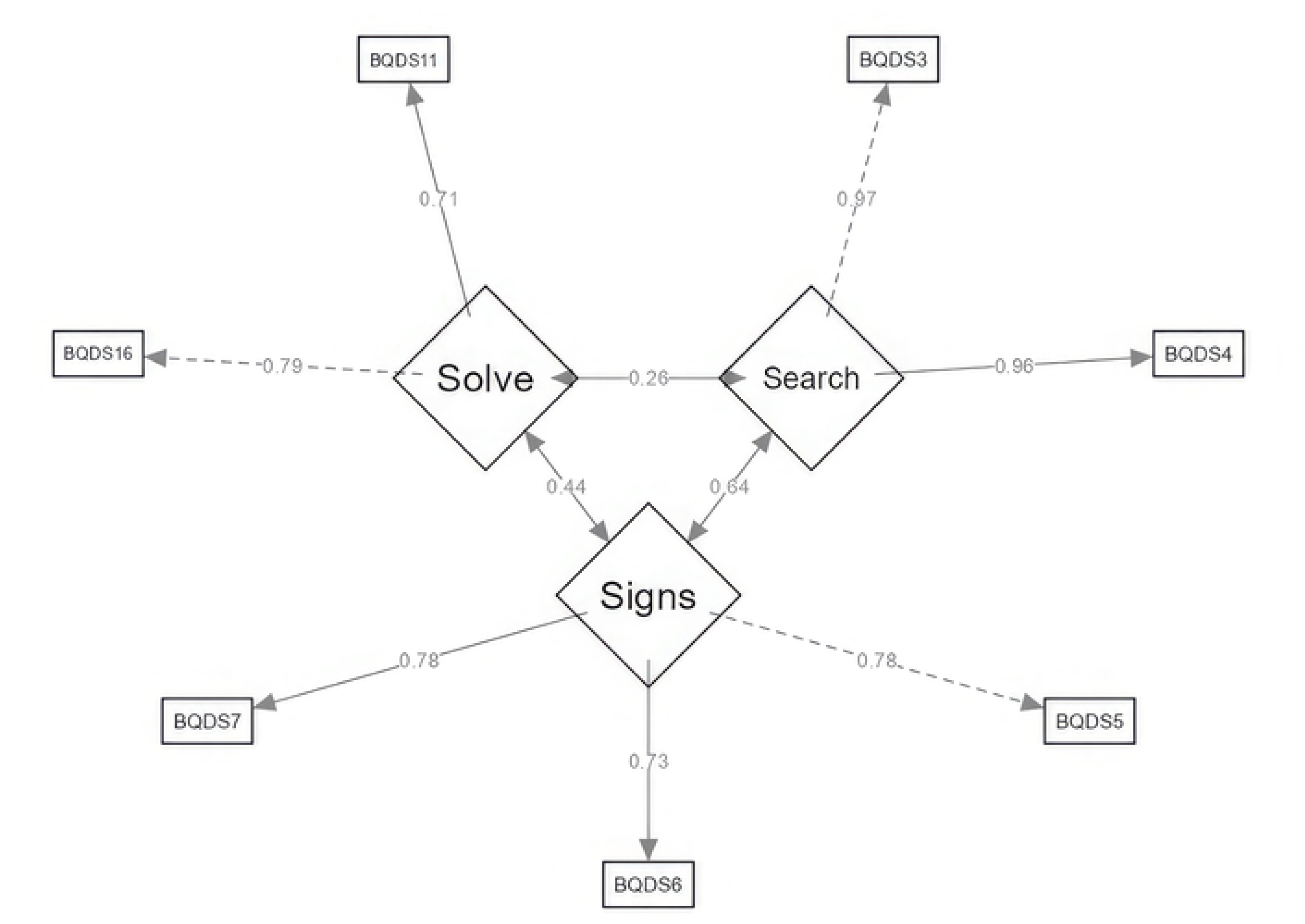
Path diagram showing factor loading of the three-factor, seven-item version of the betel quid dependency scale.

The distribution of the indicators in the proposed model was multivariate nonnormal since Mardia’s multivariate skewness and kurtosis statistics were high enough to reveal p-values larger than 0.05 [54]. In the next step, confirmatory factor analysis (CFA) via weighted least squares estimation (WLSMV) was performed to test the hypothesis that a connection exists between the observable variables and their underlying latent concept (s). Standardized factor loading and fit indices were used to evaluate structural and cultural validity and determine whether the proposed factorial structure was consistent with the population of Bangladesh. To investigate the theoretical foundations of the shorter form of the BQDS, we employed the three subscales from the EFA (search, signs, solve) in the CFA. Based on the data displayed in the above figure, the measurement model of this scale consists of three latent factors and seven observable variables. All seven items have factor loadings greater than the standard threshold of 0.60 [Fig.2].

The proposed three-factor model also met the “good fit” measure index (fit indices and fit measures) requirements, with suggested cut-off* values of GFI = 0.99 > 0.95*, NFI = 0.98 > 0.95*, TLI = 0.98 > 0.95*, CFI = 0.99 > 0.95*, RFI = 0.97 = ∼ 1*, IFI = 0.99 > 0.95*, PNFI = 0.51 > 0.50*, RMSEA = 0.06 within the acceptable values of.05 to 0.08, and SRMR = 0.03. < 0.08 [55]. These findings indicate that the three-factor Betel quid dependency scale, which holds seven items, has acceptable structural and cultural validity in the Bangladeshi population [Table 2 and Table 3].

**Table 2.**
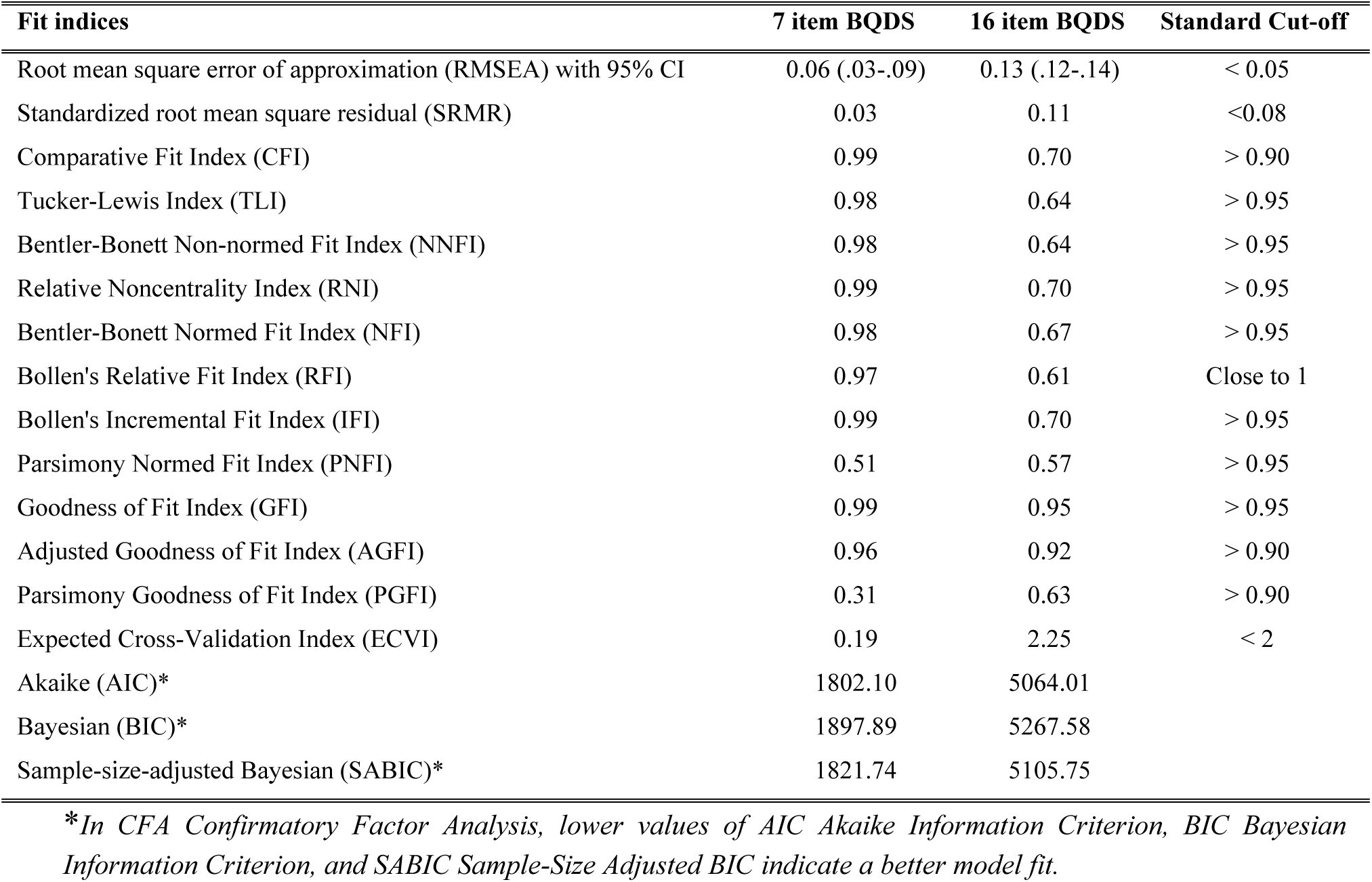
Comparison of CFA fit indices & matrices between the shorter version BQDS (7 items) vs the original BQDS (16 items)

| Fit indices | 7 item BQDS | 16 item BQDS | Standard Cut-off |
| --- | --- | --- | --- |
| Root mean square error of approximation (RMSEA) with 95% CI | 0.06 (.03-.09) | 0.13 (.12-.14) | < 0.05 |
| Standardized root mean square residual (SRMR) | 0.03 | 0.11 | <0.08 |
| Comparative Fit Index (CFI) | 0.99 | 0.70 | > 0.90 |
| Tucker-Lewis Index (TLI) | 0.98 | 0.64 | > 0.95 |
| Bentler-Bonett Non-normed Fit Index (NNFI) | 0.98 | 0.64 | > 0.95 |
| Relative Noncentrality Index (RNI) | 0.99 | 0.70 | > 0.95 |
| Bentler-Bonett Normed Fit Index (NFI) | 0.98 | 0.67 | > 0.95 |
| Bollen's Relative Fit Index (RFI) | 0.97 | 0.61 | Close to 1 |
| Bollen's Incremental Fit Index (IFI) | 0.99 | 0.70 | > 0.95 |
| Parsimony Normed Fit Index (PNFI) | 0.51 | 0.57 | > 0.95 |
| Goodness of Fit Index (GFI) | 0.99 | 0.95 | > 0.95 |
| Adjusted Goodness of Fit Index (AGFI) | 0.96 | 0.92 | > 0.90 |
| Parsimony Goodness of Fit Index (PGFI) | 0.31 | 0.63 | > 0.90 |
| Expected Cross-Validation Index (ECVI) | 0.19 | 2.25 | < 2 |
| Akaike (AIC)* | 1802.10 | 5064.01 |  |
| Bayesian (BIC)* | 1897.89 | 5267.58 |  |
| Sample-size-adjusted Bayesian (SABIC)* | 1821.74 | 5105.75 |  |

*\*In CFA Confirmatory Factor Analysis, lower values of AIC Akaike Information Criterion, BIC Bayesian* *Information Criterion, and SABIC Sample-Size Adjusted BIC indicate a better model fit.*

**Table 3.** Validity and reliability were compared between the original Betel Quid Dependency Scale (BQDS) (three factors, sixteen items)37 and the adapted and validated version (three factors, seven items).

| BQDS 16 items | BQDS 7 items |
| --- | --- |
| 1. Have you ever felt that you can not go on without betel quid? | x |
| 2. Have you experienced a strong craving for betel quid after you reduce or completely stop chewing betel quid? | x |
| 3. Whenever you want to chew betel quid but it is not available, would you spend a lot of time to find it? | √ Whenever you want to chew betel quid but not available, would you spend a lot of time to find it? |
| 4. Whenever you want to chew betel quid but it is not available, would you take extra steps and travel a great distance trying to buy it? | √ Whenever you want to chew betel quid but not available, would you take extra steps and travel a great distance trying to buy it? |
| 5. Have you felt agitated, irritated or anxious after you reduce or completely stop chewing betel quid? | √ Have you felt agitated, irritated or anxious after you reduce or completely stop chewing betel quid? |
| 6. Have you experienced difficulty in concentrating or focusing after you reduce or completely stop chewing betel quid? | √ Have you experienced difficulty in concentrating or focusing after you reduce or completely stop chewing betel quid? |
| 7. Have you experienced depression or drowsiness after you reduce or completely stop chewing betel quid? | √ Have you experienced depression or drowsiness after you reduce or completely stop chewing betel quid? |
| 8. Have you found yourself having trouble stopping chewing betel quid once you start? | x |
| 9. Have you ever chewed betel quid nonstop? | x |
| 10. Do you have a situation that amount of betel quid is gradually increased every time you chew it from the first time you experienced it? | x |
| 11. Have you felt the need to increase the amount of betel quid chewing periodically in order to achieve a pleasant or refreshing effect? | √ Have you felt the need to increase the amount of betel quid chewing periodically in order to achieve a pleasant or refreshing effect? |
| 12. Have you often found yourself chewing more betel quid than expected and/or spending more time in chewing betel quid than expected? | x |
| 13. Would you continue chewing betel quid if you find your teeth loosened or wiggled? | x |
| 14. Would you continue chewing betel quid if you have sensitive teeth (to hot or cold food)? | x |
| 15. Would you continue chewing betel quid if you experience canker sores or mouth ulcers? | x |
| 16. Have you reduced or given up any of your social, work or leisure activities because of betel quid chewing? | √ Have you reduced or given up any of your social, work or leisure activities because of betel quid chewing? |

According to the data, each latent component has AVE value of [(search = 0.93, signs = 0.58, solve = 0.54) ≥ 0.50] and CR value of [(search = 0.91, signs = 0.74, solve = 0.76) ≥ 0.70], indicating high convergent validity for the shorter version of the BQDS. The Fornell-Larcker criterion estimates discriminant validity by comparing the square root of each AVE with its correlation coefficients. The results show that the square root value of each latent component is higher than its correlation value with other factors [56] [Table 4]. The seven-item BQDS and its subscales have acceptable to high reliability, as measured by Cronbach’s alpha (α) coefficient; [ Reliability (Overall = 0.83, search = 0.96, signs = 0.80, solve = 0.69)] [57] [Table 1].

**Table 4.**
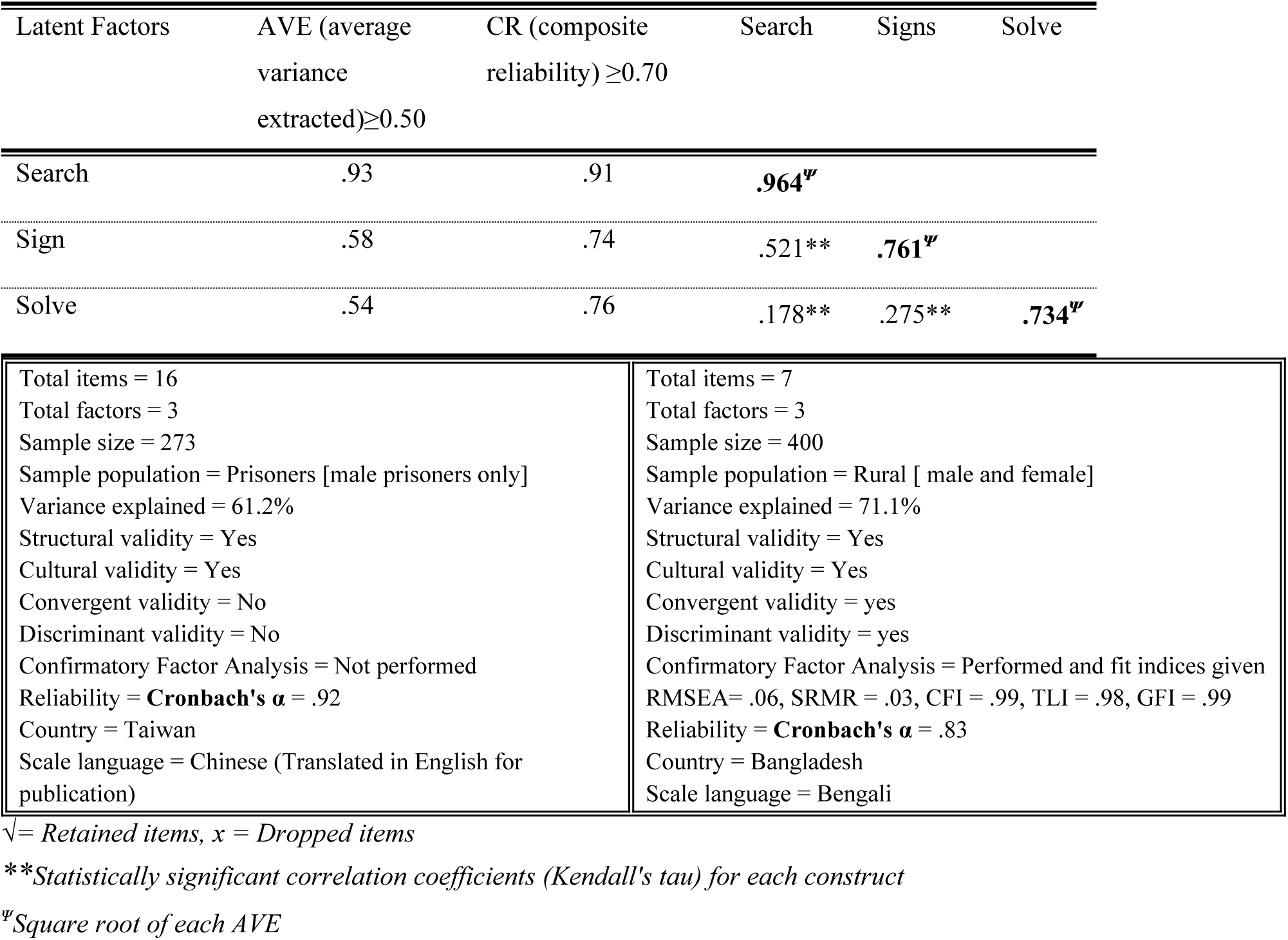
Average variance extracted (AVE), Composite reliability (CR), square root of the AVE (in bold), and correlations between constructs (off-diagonal)

| Latent Factors | AVE (average<br>variance<br>extracted) $\geq 0.50$ | CR (composite<br>reliability) $\geq 0.70$ | Search | Signs | Solve |
| --- | --- | --- | --- | --- | --- |
| Search | .93 | .91 | **.964<sup>√</sup>** |  |  |
| Sign | .58 | .74 | .521** | **.761<sup>√</sup>** |  |
| Solve | .54 | .76 | .178** | .275** | **.734<sup>√</sup>** |

|  |  |
| --- | --- |
| Total items = 16<br>Total factors = 3<br>Sample size = 273<br>Sample population = Prisoners [male prisoners only]<br>Variance explained = 61.2%<br>Structural validity = Yes<br>Cultural validity = Yes<br>Convergent validity = No<br>Discriminant validity = No<br>Confirmatory Factor Analysis = Not performed<br>Reliability = **Cronbach's $\alpha$** = .92<br>Country = Taiwan<br>Scale language = Chinese (Translated in English for publication) | Total items = 7<br>Total factors = 3<br>Sample size = 400<br>Sample population = Rural [ male and female]<br>Variance explained = 71.1%<br>Structural validity = Yes<br>Cultural validity = Yes<br>Convergent validity = yes<br>Discriminant validity = yes<br>Confirmatory Factor Analysis = Performed and fit indices given<br>RMSEA= .06, SRMR = .03, CFI = .99, TLI = .98, GFI = .99<br>Reliability = **Cronbach's $\alpha$** = .83<br>Country = Bangladesh<br>Scale language = Bengali |

√ = Retained items, x = Dropped items
*\*\*Statistically significant correlation coefficients (Kendall's tau) for each construct*
*<sup>√</sup>Square root of each AVE*

### Statistical analysis

The data were cleaned, processed, and analyzed using Statistical Package for Social Sciences (SPSS) version 23.0 and jamovi-2.6.17.0. Descriptive and inferential statistics were applied as necessary, and data were presented using tables and graphs. Exploratory factor analysis (EFA), confirmatory factor analysis (CFA), and structural equation modeling (SEM) were performed to support the adaptation and validation of the seven-item, three-factor BQ dependence scale, thereby justifying its application in assessing BQ dependency among the study participants. Applying BQDS score threshold 4, individuals were categorized into dependent and non-dependent groups [37](Tables 1 and 2, Supplementary Table 1 & 2, Fig. 2). The length of betel quid consumption was split into three categories: Group 1(less than 10 years), Group 2 (10 to 29 years), and Group 3 (30 years or more). The age at which individuals began consuming betel quid was organized into three groups (Group 1: under 20 years, Group 2: 20 to 29 years, and Group 3: 30 years and older). The daily number of betel quid consumed was divided into three categories (Group 1: less than 4 quid/day, Group 2: 4 to 6 quid/day, and Group 3: more than 6 quid/day). The reason for the BQ Chewing Scale (RBCS) and its subscales, and the intention to quit scale were analyzed as continuous variables. Nonparametric tests (χ) explored the association between socioeconomic factors and BQ dependence. Both parametric (*t-test*) and non-parametric (χ) methods were employed to examine the relationship between BQ dependence and characteristics of BQ users, such as personal habits, motivations for using BQ, usage patterns, and intention to quit BQ. To identify significant predictors for BQ dependence while adjusting for confounders, all explanatory variables that demonstrated a significant association in univariate and bivariate analyses (p < 0.05), were included in a binary logistic regression model [Enter method]. Phi (ϕ), Cramér’s V, and Cohen’s d were reported to quantify the strength of relationships between variables. The logistic regression model was assessed for goodness of fit, multicollinearity, explained variability, and overall classification accuracy. The predicted probabilities were represented by beta coefficients and adjusted odds ratios (AOR), alongside their crude odds ratios, with a 95% confidence interval and p-values. Structural equation modeling and a decision tree were developed to support, explain, and illustrate the hierarchical relationships among predictors of BQ dependency in the respondents.

## Results

More than half of the total participants, 56% (n = 224), were females, while 44% (n = 177) were males. The average age of female respondents was 41.21 ± 10.76 years, while the average age for males was 43.47 ± 11.74 years. More than 98% (n = 396) of the respondents were Muslims. Data revealed more than 50% (n = 219) of the sample population were illiterate, and about 11% (n = 45) were widows/widowers. In terms of occupation, homemakers topped the list with 50%, followed by farmers 18% (n = 72), business people 8.5% (n = 34), service 6.5% (n = 26), rickshaw puller 5.5% (n = 22), and day laborer 5.3% (n = 21). Approximately one-fourth, 25.8% (n = 103) of betel quid consumers began chewing at 18 years, and nearly half of the respondents, 48% (n = 192), had consumed betel quid more than six times per day. Fifty percent of the sample reported spending more than 500 TK per month on BQ, and 60% (n = 245) had consumed it for more than ten years. Among the 400 participants, 89.25% (n = 357) reported using tobacco in various forms along with their betel quid, while 10.75% (n = 43) did not. Of those who used tobacco, 70.25% (n = 281) exclusively used smokeless tobacco with their betel quid,6.75% (n = 27) only smoked without using smokeless tobacco, and 12.25% (n = 49) utilized both forms of tobacco. Data indicates that no female respondents had a smoking habit [Table 5]. Females had higher average scores for “betel quid dependence” and “reason for betel quid chewing” than males; [3.80 (SD = 2.08) vs 2.49 (SD = 2.12)] and [13.74 (SD = 4.16) vs 12.16 (SD = 4.38)] respectively. On the other hand, males had a higher mean” Betel quid quit” score than females [17.27 (SD = 4.17) vs 16.74 (SD = 4.49)]. [Not shown in table]. According to data, betel quid dependence was present in 48.5% (*n* = 194) of all participants. In examining the connection between sociodemographic factors and betel quid dependency, a statistically significant association was found with gender, education, and occupation (*p* < 0.001) [Table 5].

**Table 5.** Distribution and the relationship between Betel quid dependency and sociodemographic characteristics (*N* = 400)

| Characteristics | Categories | Not<br>Dependent | Betel quid<br>Dependent | Total | $\chi^2(df)$ | p-value | Phi/<br>Cramér's V |
| --- | --- | --- | --- | --- | --- | --- | --- |
|  |  | N (%) | N (%) | N (%) |  |  |  |
| Sex of the respondent | Male | 118 (67.0) | 58 (33.0) | 176 (100.0) | 29.31 (1) | 0.000<sup>ψ</sup> | 0.28* |
|  | Female | 88 (39.3) | 136 (60.7) | 224 (100.0) |  |  |  |
| Age of the respondent | less than 35 years | 58 (52.7) | 52 (47.3) | 110 (100) | 2.45(2) | 0.294 | 0.08** |
|  | 35-49 years | 79 (55.6) | 63(44.4) | 142 (100) |  |  |  |
|  | 50 years and above | 69 (46.6) | 79 (53.4) | 148 (100) |  |  |  |
| Education of the respondent | No formal education | 95 (43.4) | 124 (56.6) | 219(100) | 20.19(2) | 0.000<sup>ψ</sup> | 0.23** |
|  | Below Secondary | 63 (53.8) | 54 (46.2) | 117(100) |  |  |  |
|  | Secondary and higher | 48 (75) | 16 (25) | 64(100) |  |  |  |
| Occupation of the respondent | Unemployed | 79 (37.1) | 134 (62.9) | 213 (100) | 39.77 (2) | 0.000<sup>ψ</sup> | 0.32** |
|  | Day laborers | 77 (64.2) | 43 (35.8) | 120 (100) |  |  |  |
|  | Service | 50 (74.6) | 17 (25.4) | 67 (100) |  |  |  |
| The income of the respondent in Taka | Below 5000 Taka | 28 (48.3) | 30 (51.7) | 58(100) | 0.45 (3) | 0.930 | 0.03** |
|  | 5000 taka- 9000 Taka | 88 (51.8) | 82 (48.2) | 170(100) |  |  |  |
|  | 10000 taka- 14000 Taka | 51 (53.7) | 44 (46.3) | 95(100) |  |  |  |
|  | Above 15000 Taka | 39 (50.6) | 38 (49.4) | 77(100) |  |  |  |
| Occupation of the head of the household | Unemployed | 5 (41.7) | 7 (58.3) | 12 (100) | 2.41 (2) | 0.300 | 0.10** |
|  | Day laborers | 47 (34.3) | 90 (65.7) | 137 (100) |  |  |  |
|  | Service | 35 (44.9) | 43 (55.1) | 78 (100) |  |  |  |
| Type of family | Nuclear family | 129 (51.0) | 124 (49.0) | 253 (100) | 0.07 (1) | 0.788 | 0.01* |
|  | Join family | 77 (52.4) | 70 (47.6) | 147 (100) |  |  |  |
| Marital status | Single | 3 (60.0) | 2(40.0) | 5 (100) | 1.82 (2) | 0.403 | 0.07** |
|  | Married | 183 (52.6) | 165(47.4) | 348 (100) |  |  |  |
|  | Widow/widower/separated | 20 (42.6) | 27(57.4) | 47 (100) |  |  |  |

Characteristics associated with betel quid usage, such as whether family members chew betel quid, the age at which one starts chewing, the frequency of use, the combination of betel quid with any tobacco, the total duration of betel quid consumption, and the monthly spending on betel quid, all showed a significant relationship with dependence on betel quid. The motivations for betel quid (BQ) consumption, categorized into three key components—Reinforcement, Socio-cultural influences, and Stimulation—were all found to be statistically significantly associated with BQ dependence, with “Stimulation” exhibiting the strongest effect. Furthermore, the intention to quit BQ use demonstrated a significant inverse association with dependence (p < 0.001) [Table 6].

**Table 6.**
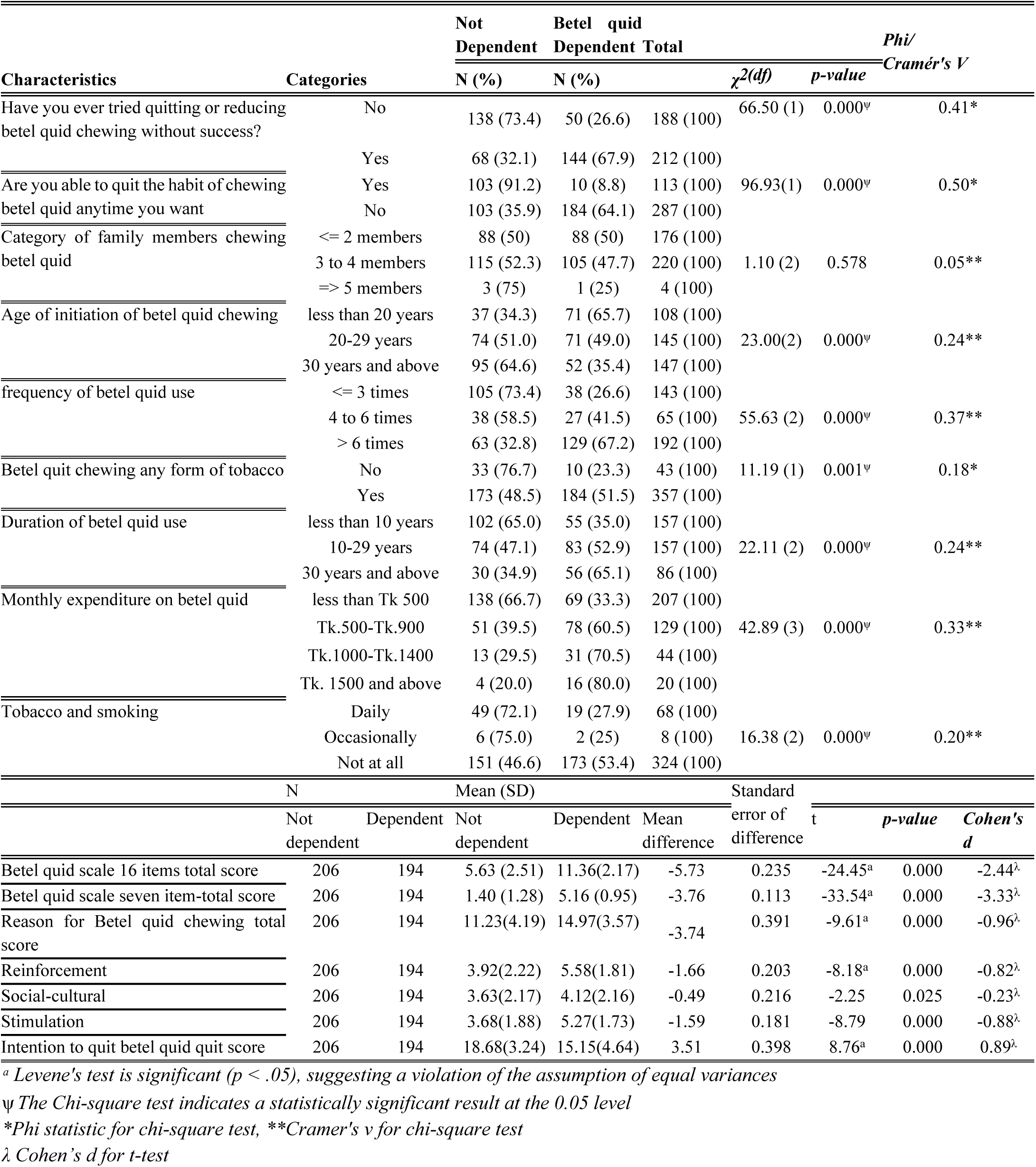
Betel quid consumption characteristics, reason for chewing, cessation behaviors, and betel quid dependence [*N* = 400].

### Binary logistic regression [assumption and model fit]

To determine the key predictors of BQ dependency while accounting for confounders, variables that showed statistical significance in the initial analyses were incorporated into a binary logistic regression model. The main explanatory variables were organized into multiple regression model blocks and analyzed using the enter method. The multicollinearity assumptions of the model variables were assessed by examining the variance inflation factors (VIFs), which ranged from 1.01 to 2.1 (All values being less than 4). Tolerances were between 0.44 and 0.99 (All greater than 0.25). The Hosmer–Lemeshow Goodness of Fit Test was conducted to evaluate the model’s overall fit, resulting in a chi-square value of 10.45 with a significance level of 0.23. The model demonstrates strong support for fit since this value is greater than 0.05. Furthermore, the McFadden, Cox & Snell, and Nagelkerke R-squared values were 0.45, 0.45, and 0.61, respectively. These values suggest that the predictor variables in this model explain a significant proportion of the variance in the dependent variable, BQ dependency. The model occupied 90% of the area under the curve and accurately identified 91% of respondents as betel quid-dependent vs 74% as non-dependent, with an overall accuracy of 84% [Figure not shown].

### Predictors for betel quid dependency, adjusting for confounders

According to the model, respondents with higher education levels were about five times less likely to become dependent on BQ compared to those with no formal education (Adjusted Odds Ratio [AOR] = 0.176; 95% Confidence Interval [CI] = 0.04 to 0.80, p = 0.02). Additionally, Day laborers had approximately thirteen times more chance of developing betel quid dependence than unemployed individuals (AOR = 12.6; 95% CI = 1.23 to 120.87, p = 0.03). The data revealed that respondents who spent more money on BQ purchases were five times more likely to have BQ dependence (AOR = 5.26; 95% CI = 1.32 to 20.87, p = 0.02). Furthermore, individuals who consumed tobacco alongside BQ were twenty times more prone to developing dependency (AOR = 19.95; 95% CI = 2.34 to 170.40, p =0.006). Respondents who had previously attempted to quit chewing BQ but had not succeeded were nearly five times more likely to develop a dependency on it (AOR = 4.73; 95% CI = 1.74 to 12.92, p = 0.002). The reasons for chewing BQ positively predicted dependency [(b = 0.14; Standard Error [S.E.] = 0.07, Wald = 3.92, p = 0.05), (AOR = 1.15; 95% CI = 1.0 to 1.32)], whereas the intention to quit had an inverse impact on BQ dependence [(b = − 0.16; S.E. = 0.06, Wald = 6.51, p = 0.01), (AOR = 0.85; 95% CI = 0.75 to 0.96) [Table 7].

**Table 7:**
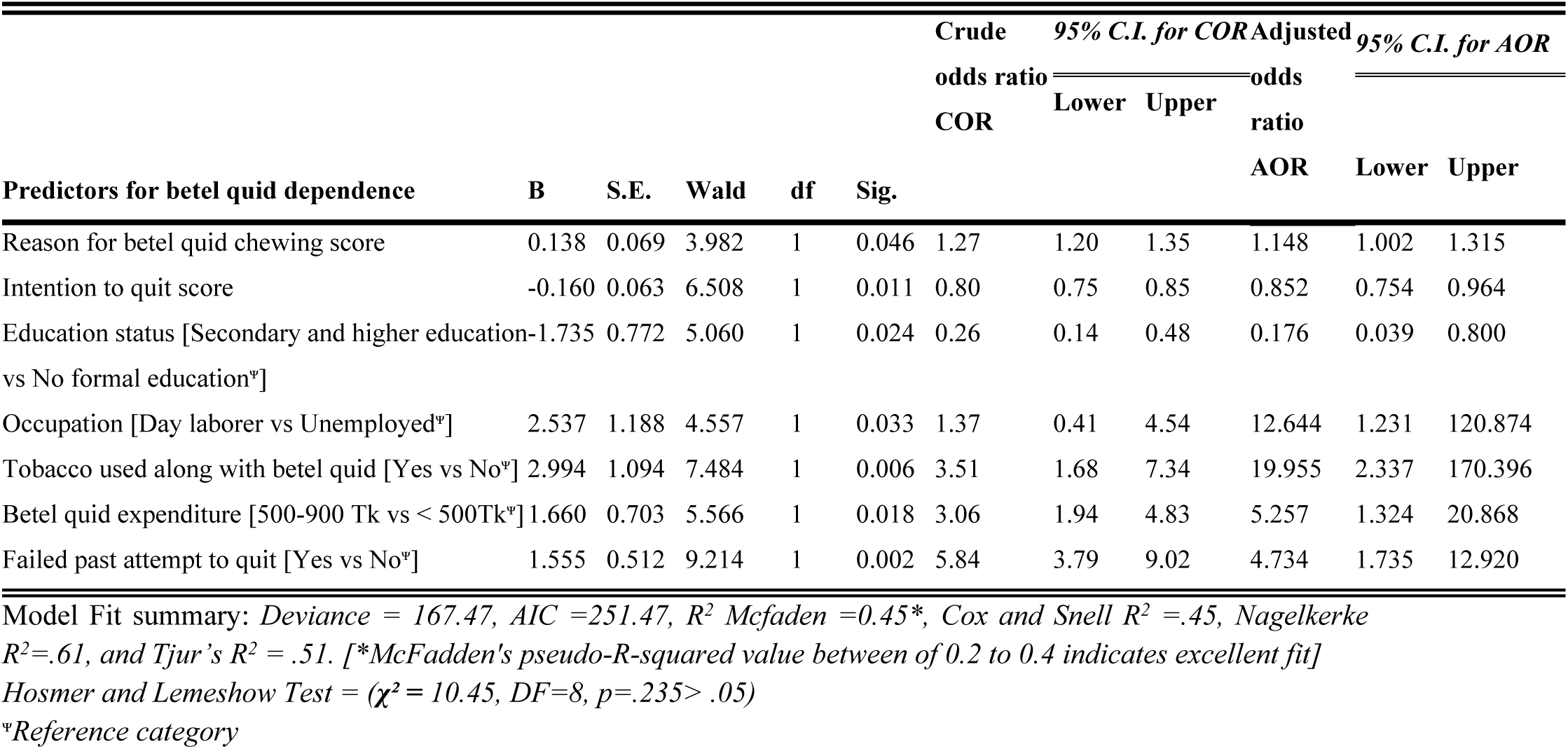
Predictors for betel quid dependence.

The structural equation modeling, represented in the path diagram, supported and highlighted the inference derived from the binary logistic regression model, conforming to the hypothesized framework of variables influencing betel quid dependence [Figs. 1 and 3].

**Fig. 3.**
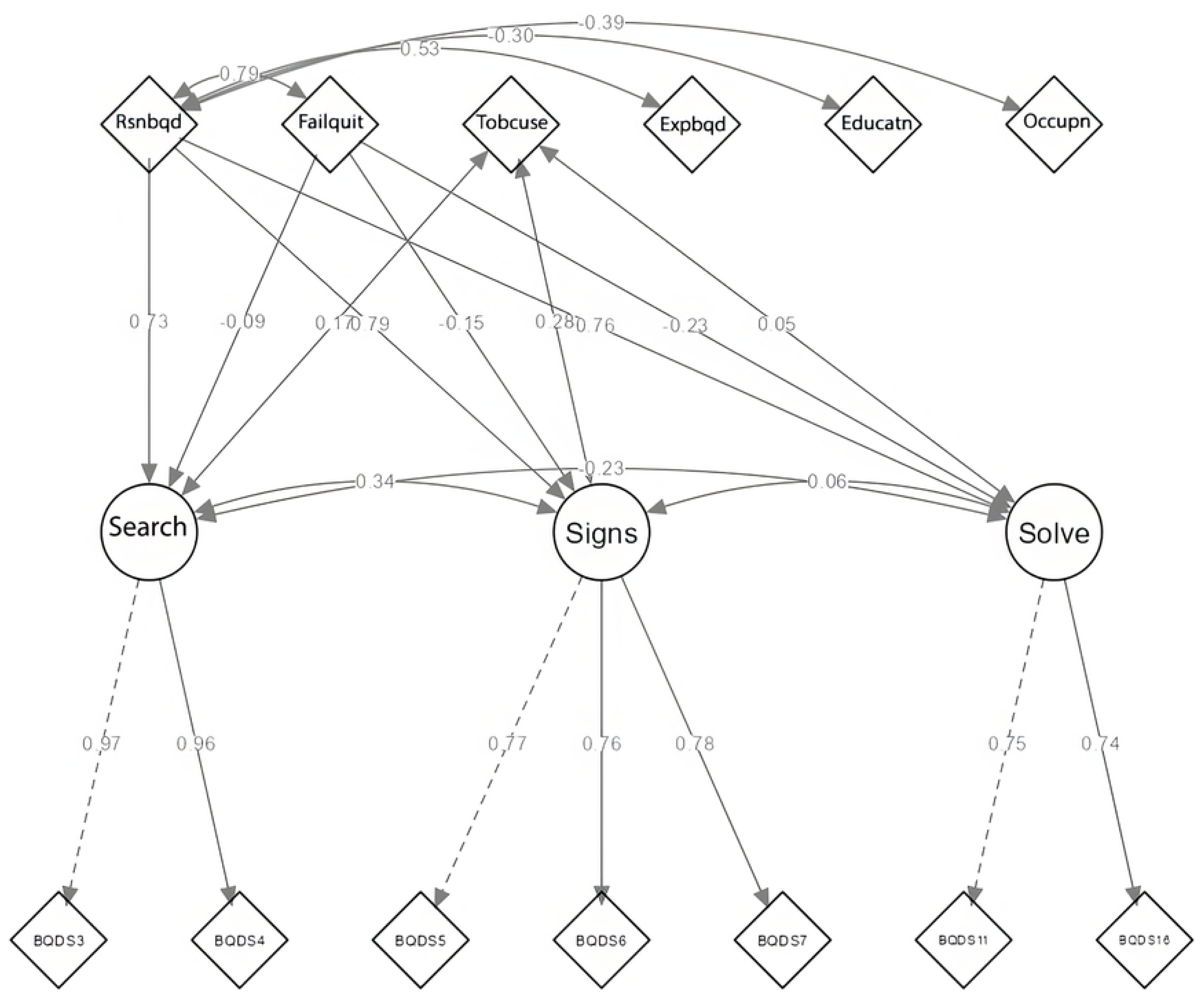
The structural equation model depicts the association between adapted BQDS subscales (search, signs, and solve) and other critical independent factors, such as the reason for betel quid chewing, failed attempt to quit, tobacco habits, expenditure for betel quid consumption, and sociodemographic characteristics. *“Failquit”* = *Failed past attempt to quit, “Rsnbqd”* = *Reason for betel quid chewing, “Tobuse”* = *Tobacco used along with betel quid, “Expbqd”* = *Betel quid expenditure, “Educatn”* = *Education status, “Occupn”* = *Occupation of the Household*.

Researchers constructed a decision tree alongside logistic regression to validate its findings, explain interactions, and provide actionable insights in a logical arrangement. This combined approach ensured both statistical rigor and practical relevance in addressing BQ dependence. The decision tree not only categorized individuals into dependent and non-dependent groups but also illustrated how different predictors interact, providing crucial insights into the hierarchical importance of factors influencing BQ dependency. Both models identified reasons for chewing BQ, the intention to quit, educational level, failed quit attempts, BQ with tobacco, and BQ expenditure as significant predictors [Fig. 4].

**Figure 4.**
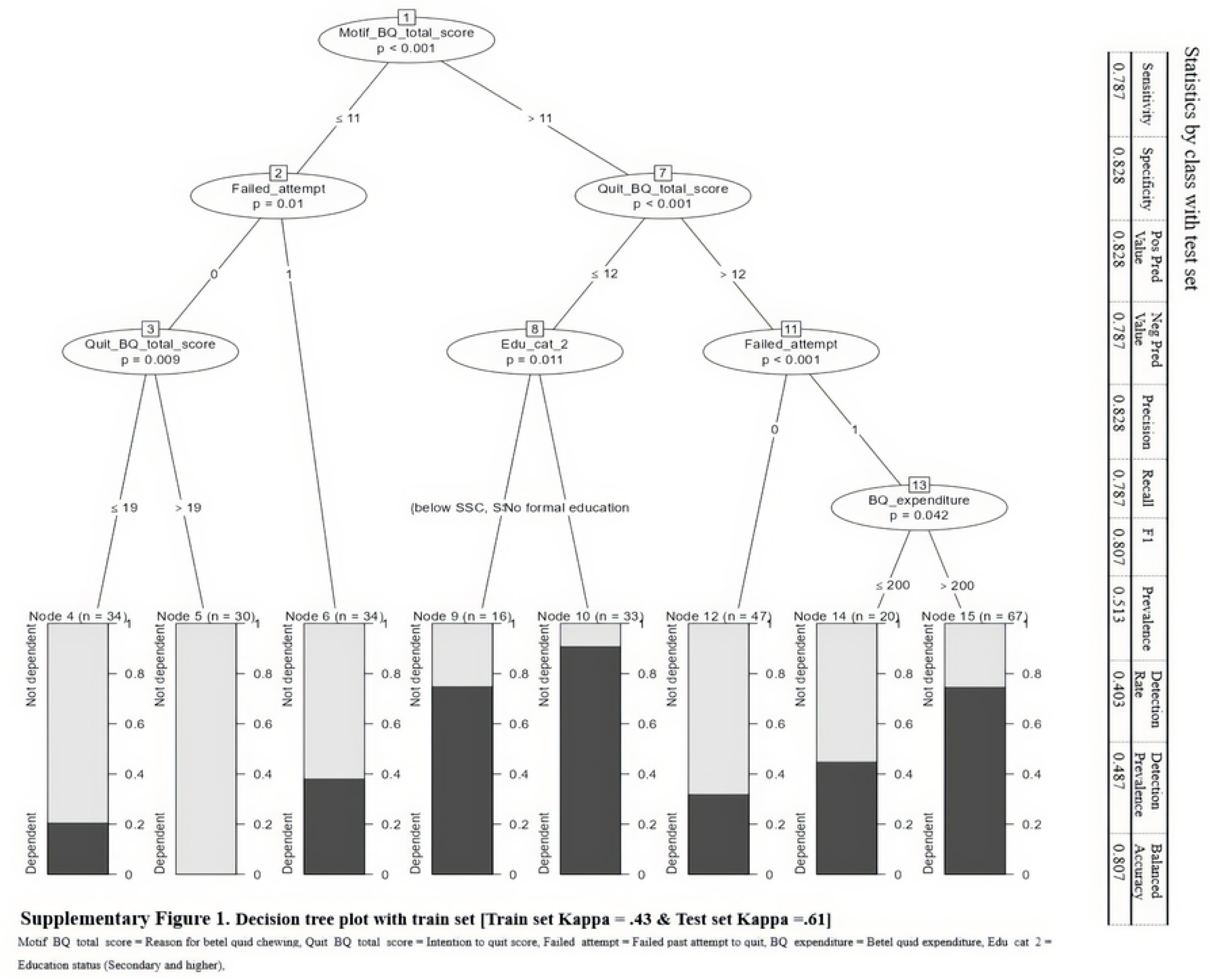
The decision tree depicted the interactions among various predictors and highlighted the hierarchical significance of factors contributing to BQ dependency.

## Discussion

This cross-sectional study was conducted in Bhaluka Upazila, a rural subdistrict of Mymensingh, Bangladesh, to assess the prevalence of betel quid (BQ) dependence and the associated influencing factors among users. The study examined various sociodemographic characteristics, patterns of BQ consumption, tobacco use behaviors, and three validated behavioral measures related to BQ use: motivations for chewing BQ, intention to quit, and level of dependence. To assess BQ dependence, we adopted and validated a shortened version of the Betel Quid Dependence Scale (BQDS), formerly developed by Lee et al. [37] and subsequently replicated in Guam [38]. The original BQDS consists of three factors and sixteen items, demonstrating strong internal consistency and construct validity. However, its initial development and validation were limited to male prisoners in Taiwan with a history of BQ use. Moreover, the scale was initially formulated in Chinese and later translated into English without evaluating its psychometric properties in English or other languages. Before being used in this study, the shortened seven-item BQDS was assessed for structural and cultural validity, convergent and discriminant validity, and reliability. This validation process was performed to justify the appropriateness of the shortened BQDS for Bengali-speaking male and female BQ chewers in rural Bangladesh, ensuring its relevance for clinical and social research applications within Bangladeshi and Bengali-speaking populations (Supplementary Table 1).

In this study, participants began chewing betel quid (BQ) at ages ranging from as young as 10 to as old as 58, with an average starting age of 26.35 years (SD = 11.11). Notably, 25% of individuals had tried BQ by the age of 18. This finding aligns with an Assamese study that indicated the initiation age for areca nut chewing typically falls between 12 and 20 years. [58]. Likewise, Lee et al. reported that male inmates in Taiwan started chewing BQ at an average age of 18.21 years [59], while Murphy et al. found that Chamorro and non-Chamorro Micronesians began areca nut consumption at around 17.77 years [60]. Almost half of the respondents in this study spent more than 500 Bangladeshi Taka (Tk) per month on BQ, with an average monthly cost of 548 Tk (SD = 412.67). A prior study revealed that BQ users spent approximately 2.29 Tk daily (equivalent to around 600 Tk monthly), with expenditure levels influenced by factors such as marital status, educational background, religious affiliation, and employment status [4]. Regarding usage patterns, participants reported chewing an average of eight BQ per day, with a mean duration of use of 15.82 years (SD = 12.36). Several other studies found that regular users chewed BQ around 5.15 times daily, with differences depending on their occupation, religion, age, and marital status [4,61]. Similarly, an Indian study showed that people chewed BQ between 3 and 6 times a day, with the duration of use ranging from 10 to 30 years, averaging 18 years [58].

In this study, female participants had higher mean scores for both the “reasons for chewing betel quid (BQ)” and “BQ dependence” compared to males. Conversely, males had a higher average score for the “intention to quit BQ” than females. Similar findings have been reported in Malaysia and Pakistan, where women were more likely to start chewing BQ and less likely to quit [62,63]. Approximately three-fourths of the respondents (*n*= 287) stated their inability to discontinue betel quid. Since they began consuming it, more than half of the sample (*n*= 212) desired to quit or reduce their betel quid consumption. Only a tenth of the participants (*n=42*) attempted to stop chewing more than twice. The data revealed that betel quid dependence was present in 48.5% of all participants (*n=194*), with female users being nearly twice as likely to be dependent on betel quid as male users. A study on oral cancer and betel quid dependency across six Asian ethnicities found an addiction frequency ranging from 0.8% to 46.3%. Over 40.5% of current chewers reported abuse, especially among Nepalese and Southeast Asians [12,16].

Men showed a higher prevalence of BQ dependency in East Asia, while women dominated in Southeast Asia [12]. The current study reports that females have a higher dependency score and a greater proportion of betel quid (BQ) dependency. This trend may be influenced by various demographic and behavioral factors, such as education level, tobacco use in conjunction with BQ (primarily smokeless), failed attempts to quit BQ, reasons for chewing BQ, and intentions to quit BQ. Tobacco-laced betel quid is prevalent in lower-income groups, whereas non-tobacco use is more common among those with moderate education [42]. Among BQ-tobacco users, 57% are female, primarily using smokeless tobacco, while 43% are male, who mainly smoke or use both forms. Of those who attempted to quit betel quid,71% of females were unsuccessful compared to 29% of males. These findings are consistent with India’s Global Adult Tobacco Survey (GATS). In India, the use of betel quid among young males decreased by 40% across two surveys when it was consumed with tobacco, while the usage among females remained unchanged [42].

In our study, respondents with higher education levels were about five times less likely to become dependent on betel quid than respondents with no formal education. Daily laborers were about thirteen times more likely to develop betel quid dependency than those who were jobless. According to data, respondents who spent more money on betel quid purchases were five times more likely to acquire a dependence on betel quid. Research indicates that betel quid chewing is particularly common among women, older individuals, tobacco smokers, and those with lower socioeconomic status, including people with limited education, lower incomes, and no land ownership [3,6,41,42,63,64].

In the current study, individuals who used tobacco with betel quid were found to be twenty times more likely to become dependent on it. Additionally, those who had previously attempted to quit chewing betel quid but were unsuccessful were nearly five times more likely to develop an addiction to the habit. According to Ghani et al., Indians with a history of smoking are more prone to acquire a quid chewing habit, and quitting is less probable among those who incorporate areca nut and tobacco in their quid [61,64]. In another study, concurrent use of tobacco and betel quid has also been shown to be a major predictor of betel quid addiction [25,61], and persistent and compulsive BQ use contributes to the progression of Oral Squamous Cell Carcinoma [32]. Findings indicate that the composite score for “Reason for betel quid chewing,” which aggregates the scores of “stimulation,” “reinforcement,” and “socio-cultural” themes, suggests probable dependency. However, the “Intent to quit” presents an opposing impact. A comprehensive systematic review and a study conducted in Guam support this behavioral trend. [33,34]. Among self-identified betel-quid chewers in Guam and adolescent schoolchildren in Karachi, “stimulation” emerged as the primary motivation for chewing, followed by “reinforcement” and “social/cultural” influences [34,62,63]. A recent systematic review found that habitual BQ users face challenges quitting due to addiction, withdrawal symptoms, and ingrained behavior. In Southeast Asia, social chewers often resist quitting, as they see areca nut as a key part of their cultural and social identity, with peer influence reinforcing the habit [31,62,63,64]. These findings align closely with the results of our study (Table 4).

### Limitations

This study identified several limitations that may impact the validity of the findings. First, relying on an interview-administered questionnaire as the primary data collection method limited the depth of insights. Qualitative methods, such as open-ended interviews or focus groups, could have provided richer perspectives on betel quid dependence. The sample size for this study was 400, which narrowly fulfilled the sampling adequacy criterion for the logistic regression model specified by Hosmer and Lemeshow [65]. Since participants were drawn from only one subdistrict through simple random sampling, the findings may not be generalizable to the broader population of Bangladesh; cultural and socio-economic factors can vary significantly across regions. The cross-sectional approach of the study limits our ability to explore temporal relationships. This prevents us from establishing causal inferences about the relationships between betel quid dependence and sociodemographic factors, the reasons for chewing betel quid, and quitting behavior. As a result, we cannot determine whether specific patterns of betel quid use lead to dependence or if dependence leads to these patterns of use. To gain a more thorough and nuanced understanding of BQ dependence dynamics, studies should utilize representative samples and adopt longitudinal study designs.

## Conclusion

An adapted shortened version of the Betel Quid Dependency Scale showed satisfactory structural and cultural validity and acceptable convergent and discriminant validity, including good reliability. This study elucidates the effects of educational attainment, occupational status, tobacco use, quitting behavior, and unsuccessful quit attempts on BQ dependence and the behavioral motivations behind BQ consumption. Targeted interventions such as health education campaigns, behavioral and social support initiatives, and strengthened anti-tobacco policies have significant potential to raise awareness, reduce betel quid consumption, and alleviate the rural health burden caused by BQ dependency.

## Data Availability

The data that support the findings of this study are openly available in figshare at . https://doi.org/10.6084/m9.figshare.27101053.v3 and URL https://figshare.com/articles/dataset/_b_Betel_Quid_Dependence_in_Rural_Adults_Bangladesh_Context_b_/27101053/3?file=50312721

## Abbreviations

BQ: Betel quid
BQDS: Betel quid dependency scale
RBCS: Reason for betel quid chewing scale
EFA: Exploratory factor analysis
CFA: Confirmatory factor analysis
SEM: Structural equation modeling
AVE: Average variance extracted,
CR: Composite reliability.

## Acknowledgements

Community Medicine & Public Health Department of Patuakhali Medical College, Bhaluka Upazila administration, Mymensingh.

